# Myeloid subpopulations correlate with responsiveness to therapy in a prospective, longitudinal study on RA patients

**DOI:** 10.64898/2026.11.02.26367113

**Authors:** Mohammad D. Khan, Matthew Dapas, Salina T. Dominguez, Tyler Therron, Kathleen Aren, Mary Carns, Hadijat M. Makinde, Yidan Wang, Samuel D. Dowling, Richard M. Pope, Arthur M. Mandelin, Eric M. Ruderman, Joan M. Bathon, Laura B. Geraldino-Pardilla, Lutfiyya N. Mohammed, Anh Chung, John F. Seagrist, Ruth Misha, Yvonne C. Lee, Carla M. Cuda, Deborah R. Winter, Harris Perlman

## Abstract

**Background:** Rheumatoid arthritis (RA) is an autoimmune disease associated with the destruction of cartilage and bone, increased risk of cardiovascular disease and a higher mortality rate. Despite the many therapies that exist for RA, there is little information to guide selection of the most effective treatment for an individual patient. Ultrasound-guided synovial biopsies have created an opportunity to investigate the target tissue. While a number of studies have characterized the stromal and immune cell populations using single-cell technology on synovial biopsy samples, here we performed a longitudinal study, where we collected comprehensive clinical data over a one-year period on each patient in the process of changing their therapy.

**Methods:** Samples from minimally invasive, ultrasound-guided synovial biopsies were collected from RA patients at baseline prior to start of a new therapy and 4-6-week follow-up under IRB-approved protocols with informed consent. Comprehensive clinical data from each appointment were recorded into a shared REDCap database. Failure to respond to therapy was based on clinical disease activity index (CDAI) for RA after 12-18 weeks post therapy. Biopsied samples were dissociated, enriched for CD45⁺ or CD45⁺CD11b⁺ cells, and processed on the 10X Genomics Chromium platform to generate libraries for single-cell RNA sequencing with cell-surface protein detection (CITE-seq). Spatial transcriptomics were performed on formalin-fixed paraffin embedded sections analyzed with 10X Xenium and NanoString CosMx.

**Results:** We performed CITE-seq analysis on ∼38,000 CD45+ synovial cells isolated from 17 patients who failed a conventional or biologic DMARD and planned to change therapy. We then subclustered on the myeloid population to reveal 12 myeloid subpopulations that consist of macrophages (n = 8), Osteoclast-like cells, DCs (n = 2) and proliferating cells. The proportion of cDCs was significantly higher in responders, while the proportion of macrophages was significantly lower in responders. The proportion of *ISG15^+^*and *THBS1^+^*macrophages were enriched in the non-responder patients. Spatial transcriptomics revealed that *ISG15^+^* macrophages were preferentially localized to the synovial lining, while the *THBS1^+^* macrophages are likely interacting with lymphoid cells within the sublining tissue.

**Conclusions:** These data are among the first to prospectively profile macrophage populations as predictors of response to therapy. Future studies are required to verify whether myeloid subpopulation associations hold across specific therapies.

## INTRODUCTION

Rheumatoid arthritis (RA) is a chronic autoimmune disease that results in cartilage and bone destruction and increased morbidity and mortality. Currently, there are a plethora of therapies available for patients, yet approximately 40-50% of patients fail to achieve a robust response to initial therapy [1]. Moreover, around 25% of patients are refractory to multiple therapeutics [1]. Decisions for treatment continue to rely on trial and error as there are no prognostic indicators that predict successful response. Previous studies have shown that the number of sublining macrophages relates to disease activity and is an independent risk factor for severe progression of bone erosion[2–4]. Furthermore, change in the number of sublining macrophages is currently the only known biomarker that is associated with response to therapy [5, 6]. We previously showed that synovial CD206^+^ macrophages in RA patients variably express six transcriptional modules associated with therapy and disease severity [7].

Over the past several years, there have been numerous studies to identify the heterogenous nature of the synovium. The Accelerating Medicines Partnership (AMP) was the first to utilize single-cell RNA sequencing (scRNA-seq) on isolated CD14^+^ cells from the synovia of RA and osteoarthritis (OA) patients based on abundance in different patients and enriched pathways [8]. In a more extensive study using a larger dataset, AMP identified 15 subpopulations of myeloid cells including 9 macrophage subpopulations [9]. The authors identified cell-type abundance phenotypes (CTAPs) that were enriched in specific cell states, although CTAPs failed to correlate with clinical outcomes [9]. Additional studies reported ability to separate macrophage populations based on the status of anti-citrullinated protein antibody [10, 11]. Alivernini et al identified 9 different subpopulations of myeloid cells in the synovium of healthy controls and RA patients and compared the proportions of subpopulations across different classes of RA (naïve, resistant, remission) and healthy controls, respectively [12]. Hanlon et al [13] reported similar results to Alivernini et al [12] including 9 subpopulations of macrophages based on flow cytometry and scRNA-seq. More recently, Mantel et al determined that one of the macrophage sublining populations expressing *SPP1* colocalizes with fibrin [14]. While these studies were able to extrapolate the heterogenous subpopulations of macrophages and assign roles based on their transcriptional profiles, a major weakness was the studies were all retrospective or cross sectional. Prospective transcriptional studies, which include Pathobiology of Early Arthritis (PEAC) [15, 16], Rituximab Versus Tocilizumab in Anti-TNF Inadequate Responder Patients with Rheumatoid Arthritis (R4RA) [17, 18], and Stratification of Biological Therapies for RA by Pathobiology (STRAP) [19, 20], demonstrated that histological scoring, and pathotype or endotype are not sufficient to predict therapeutic response to an individual therapy. These studies also showed that transcriptional analysis of synovial tissue may have potential to provide predictive value for response to therapy. However, all of these studies utilized bulk tissue RNA sequencing and deconvolution methods for analysis, which together are not as robust as single cell analysis [15, 17–20].

In this study, we performed a prospective longitudinal analysis of RA patients who were either naïve to any therapy or were switching therapy. All patients underwent an ultrasound guided synovial biopsy at baseline, from which tissue was used for cellular indexing of transcriptomes and epitopes by sequencing (CITE-seq) and/or spatial transcriptomics. We observed that the proportion of macrophages were enriched in non-responders, while conventional dendritic cells (cDCs) were enriched in the responder populations. We observed the *ISG15^+^* and *THBS1^+^* macrophage subpopulations were enriched in non-responders. The *ISG15*^+^ macrophages were localized to the lining, while the *THBS1^+^* cells may be found in the sublining. These findings suggest that specific macrophage states in the synovium before treatment can predict subsequent therapeutic response.

## MATERIALS AND METHODS

The study protocol received approval from the Northwestern University Institutional Review Board, and informed consent was obtained from all participants. Adult patients aged 18-80 years with adult-onset RA met either the 1987 ACR criteria or 2010 ACR/EULAR criteria [21, 22] and had at least moderately active disease were considered as candidates for ultrasound guided synovial biopsy if they had palpable synovitis documented by clinical examination and a gray scale synovitis score of ≥2 on a 4-point scale as previously described [7]. Patients who were DMARD-naïve, defined as those who had not received any DMARD therapy but may have been treated with NSAIDs and/or steroids, as well as patients who failed conventional synthetic DMARD or biologic DMARD therapy, were recruited. All patients had at least one inflamed joint (knee, wrist, elbow, ankle). Previous studies have shown that paired wrist and knee biopsies exhibit similar histologic and transcriptional features [23–25]. At least 12 pieces of synovial tissue were retrieved, consistent with published studies [25–34]. A minimum of 8 pieces were processed for CITE-seq analysis and 4 for histology. A second biopsy was performed at 4-6 weeks following the 1^st^ synovial biopsy for a subset of patients. Clinical disease activity index (CDAI) classification of patients was divided into 4 categories of disease activity; remission ≤ 2.8, low disease activity > 2.8 to ≤ 10, moderate disease activity > 10 to ≤ 22 and high disease activity > 22 [35]. Treatment efficacy was determined at 6 and 12-18 weeks following the first biopsy as the CDAI change from baseline [35]. Patients who achieved 20% improvement in their CDAI score (CDAI-20) were considered responders, while those that failed to achieve CDAI-20 were considered non-responders. Associations between responder and non-responder were calculated using Fisher’s exact test for categorical variables and Wilcoxon rank-sum test for continuous variables. The <u>exclusion criteria</u> were (1) pregnant or lactating women, (2) active infection, (3) uncontrolled renal, hepatic, hematologic, gastrointestinal, endocrine, pulmonary, cardiac, or neurologic disease, (4) history of bleeding disorder, (5) platelet count (within one month of biopsy) less than 100 x 10^3^ cells/ µL, (6) history of infection with human immunodeficiency virus (HIV), hepatitis B, or hepatitis C, (7) documented diagnosis of SLE or seronegative spondyloarthropathy, (8) soft tissue or intra-muscular corticosteroid injections within 4 weeks prior to biopsy, (9) corticosteroid injection in joint to be biopsied within 8 weeks prior to biopsy, (10) current use of warfarin, clopidogrel, or other anticoagulant and (11) prednisone dose > 10 mg daily. Clinical data were recorded in REDCap, including demographics, disease duration, past medical history, current and prior medications, rheumatoid factor (RF) and anti-CCP status, C-reactive protein (CRP), erythrocyte sedimentation rate (ESR), Patient Global Assessment, Health Assessment Questionnaire (HAQ), Routine Assessment of Patient Index Data (RAPID3), Patient-Reported Outcome (PRO), instrument administration, Modified Health Questionnaire (mHAQ), CDAI, and DAS28.

### Tissue processing for single cell suspensions and cell sorting

Human samples were processed as previously described by us [7] and in agreement with standard operating procedures (SOPs) developed by the AMP consortium [36]. We performed scRNA-seq or CITE-seq on CD45^+^ or CD45^+^CD11b^+^ cells on the same day as the biopsy and froze the CD45^-^ cells in cryostor. Single-cell samples were run on the automated 10X Genomic Chromium platform. A portion of the partitioned sample was used to generate the ADT (surface marker) library in parallel (**Supplemental Table 1**). Prepared sample libraries were pooled, quality control (QC) by Tapestation performed by the Division of Rheumatology-Pulmonary and Critical Care Next Generation Sequencing (NGS) facility, and sequenced on an Illumina Novaseq machine. Before full sequencing, MiniSeq runs were performed to ensure the quality of samples and to estimate cell number for more accurate sequencing depth.

### Single-cell data processing and analysis

Raw single-cell RNA-sequencing data generated on 21 patient samples (17 baseline, 4 follow-up) using the 10x Genomics platform were processed with Cell Ranger (v7.0.0) using the cellranger count pipeline. Reads were aligned to the human reference genome GRCh38-3.0.0 with include-introns = false, and gene-by-cell UMI count matrices were generated. Ambient RNA contamination was corrected using SoupX (v1.6.2).

All downstream analyses were performed in R using Seurat (v5.2.1). Cells with mitochondrial transcript content greater than 15% were excluded, and putative doublets were identified and removed using DoubletFinder (v2.0.4). Following quality-control filtering and doublet exclusion, a total of 37,459 high-quality cells were retained for downstream analyses (**Supplemental Table 2**). RNA expression counts were normalized using SCTransform, with Seurat-derived S phase and G2/M phase cell cycle scores (S.Score and G2M.Score), computed from canonical cell cycle gene sets, regressed out during normalization. A total of 3,000 highly variable genes identified by SCTransform were used for principal component analysis (PCA). Sample integration was performed in PCA space using Harmony to correct for inter-sample technical variation. The Harmony-corrected embeddings were subsequently used for UMAP visualization, nearest-neighbor graph construction, and graph-based clustering using the Seurat functions RunUMAP, FindNeighbors, and FindClusters, respectively. The first 10 Harmony dimensions were used for downstream analyses, and clustering at a resolution of 0.1 identified 11 transcriptionally distinct clusters. Clusters were labelled as myeloid, T cells, B cells, Plasma cells, or mast cells based on canonical markers. For CITE-seq data, antibody-derived tag (ADT) counts were normalized independently for each sample using centered log-ratio normalization in Seurat (margin = 2). ADT measurements were used for downstream protein-level visualization and to support cluster annotation. Visualization shows log-normalized RNA expression counts. De novo marker genes for each cluster were identified using the Seurat FindAllMarkers function with only.pos = TRUE and min.pct = 0.25 (**Supplemental Tables 4-5**).

### Myeloid cell sub-clustering and label transfer

For myeloid sub-clustering, cells corresponding to the myeloid compartment were first extracted from the full dataset. To ensure robust annotations, we chose 5 representative samples containing more than 300 myeloid cells and having a median UMI count greater than 2000 as the reference set for myeloid sub-clustering. RNA expression data were re-normalized using SCTransform for each sample individually, using the default number of 3,000 highly variable genes, and Harmony was used for integration. The first 10 dimensions were used for downstream analysis, and clustering was performed at a resolution of 0.45. Sub-cluster identities were assigned on the basis of canonical marker genes together with *de novo* cluster genes (defined as above).

Following annotation of the reference myeloid subset, subcluster labels were transferred to myeloid cells from all other samples using the Seurat reference-mapping workflow. Briefly, myeloid cells from all samples were independently normalized using SCTransform and transfer anchors were identified between the annotated reference object and the query cells using Seurat’s FindTransferAnchors with normalization.method = “SCT”. Predicted subcluster labels were then assigned using TransferData, and query cells were mapped onto the reference UMAP using MapQuery.

### Association testing of myeloid cell-type proportions

Associations between myeloid cell-type abundance and clinical response status were assessed using Mixed-effects modeling of Associations of Single Cells MASC (v0.1.0-alpha) [37]. For this analysis, only baseline samples were analyzed (responder = 12, non-responder = 4, 1 patient excluded due to lack of follow-up). Only cell types represented by at least five cells in each comparison group (responder vs. non-responder) were retained. A logistic mixed-effects model was fitted for each myeloid cell type, with response group included as the contrast of interest, age included as a fixed effect, and sample identity included as a random effect. Likelihood ratio testing was used to assess the significance of the response term, and odds ratios with 95% confidence intervals were calculated from the fitted models. Adjusted p values were calculated using the Benjamini–Hochberg false discovery rate (FDR) method.

### Pseudobulk differential expression analysis

Differential gene expression analysis was performed using a pseudobulk strategy with edgeR (v4.0.16) [38]. For each cell population of interest, raw RNA counts were aggregated at the sample level to generate pseudobulk expression profiles. Only genes with non-zero expression in at least 50% of pseudobulk samples were retained for testing. Normalization and robust dispersion estimation were performed using edgeR. Differential expression between responders and non-responders was then tested using generalized linear models and likelihood-ratio testing. Genes were considered differentially expressed with P value < 0.05 and an absolute log_2_(fold change) > 0.5. For visualization of individual genes, pseudobulk expression was scaled to counts per million (CPM).

### Gene set enrichment analysis

Gene set enrichment analysis (GSEA) was performed on the pseudobulk differential expression results using the fgsea package (v1.31.0) [39]. All genes retained following filtering in the edgeR pseudobulk differential expression analysis were included in GSEA. Genes were ranked using a composite statistic defined as −log_10_(raw P value) × log_2_(fold change), such that both statistical significance and effect size contributed to the ranking. Hallmark gene sets, KEGG canonical pathways, and Reactome canonical pathways for Homo sapiens were obtained from the Molecular Signatures Database (MSigDB)[40] using the msigdbr package (v7.5.1). The fgsea algorithm was run with 20,000 permutations on gene sets with a minimum size of 5 genes and a maximum size of 1,000 genes. Enrichment results were summarized using normalized enrichment scores (NES), nominal P values, and Benjamini–Hochberg FDR-adjusted P values.

### Analysis of differentially expressed genes by subtype

For each differentially expressed gene from the baseline macrophage pseudobulk edgeR analysis, mean log-normalized RNA expression by subtype for each sample were averaged within responder and non-responder groups. The resulting gene-by-group expression matrices were standardized by row using Z-scores and clustered separately by K-means based on increased (K = 4) or decreased (K = 3) expression and in responders vs. non-responders. The number of clusters was selected based on the resulting expression patterns and biological interpretability of the gene groups. Cluster-level expression profiles were summarized by calculating the mean Z-score across all genes in a cluster for each macrophage subcluster and response group. Heatmaps and cluster profiles were generated using ComplexHeatmap and ggplot2, respectively. The relative contribution of individual subtypes to macrophage expression was calculated as the counts from each subtype over the total macrophage counts for the given gene in each sample; these values were then averaged across samples within responder and non-responder groups. The scaled contribution for each gene was calculated by multiplying the average subtype contribution by the average CPM across samples for each responder group.

### Statistical Analysis of Clinical Associations

Associations between myeloid cell proportions and clinical characteristics were assessed using 16 baseline RA synovial biopsy samples. Associations with clinical variables, including treatment outcomes, demographics, serology, medication history, and baseline disease characteristics, were assessed using the Wilcoxon rank-sum test for binary variables and Spearman’s rank correlation for continuous variables. Effect sizes for binary variables were calculated using rank-biserial correlation coefficients. Adjusted p-values were calculated using the Benjamini–Hochberg false discovery rate (FDR) method. Associations were visualized using heatmaps displaying the corresponding effect sizes, with nominal significance indicated at p < 0.05.

### CosMx spatial transcriptomic analysis

Spatial transcriptomic data were generated as previously described [41] using one patient on the NanoString CosMx 1,000-gene panel and obtained from NanoString output files containing the gene-by-cell expression matrix and the corresponding cell-level metadata. Initial quality-control filtering was performed by retaining cells with total RNA counts between the 5th and 98th percentiles of the dataset.

For doublet identification, the filtered dataset was first processed using SCTransform, followed by DoubletFinder with pK optimization using paramSweep, summarizeSweep, and find.pK. The expected number of doublets was adjusted for homotypic doublets using the modelHomotypic function. Following doublet removal, cells were re-normalized using SCTransform, and variable features were identified. Spatially informed clustering was then performed using BANKSY (v0.99.13). Briefly, RunBanksy was applied to the normalized RNA assay using the cellular spatial coordinates with lambda = 0.2 and k_geom = 15. Principal component analysis was then performed on the resultant BANKSY assay, which is a concatenation of the normalized gene expression matrix and a spatial neighborhood expression matrix. Harmony integration was carried out at the field-of-view (FOV) level using RunHarmony on the BANKSY PCA embeddings to account for tissue sampling variability. The Harmony-corrected BANKSY embeddings were subsequently used for UMAP generation (RunUMAP), nearest-neighbor graph construction (FindNeighbors), and graph-based clustering (FindClusters). The first 11 principal components and a clustering resolution of 0.7 were selected for downstream analyses. Cluster marker genes were identified using Seurat’s FindAllMarkers function and interpreted together with canonical markers for major synovial cell populations.

### Xenium spatial transcriptomic data processing and quality filtering

Xenium spatial transcriptomic data were generated on 11 patients using a 196-gene panel (**Supplemental Table 3**) and were processed using Xenium Ranger, and the resulting output matrices were imported into R with mols.qv.threshold = 20. To define spatially coherent tissue sections within each sample, density-based spatial clustering was performed independently for each sample using DBSCAN (eps = 100, minPts = 10) on the global x and y coordinates to identify individual tissue density clusters (TDC), roughly equating to pieces of tissue, for quality control. For each TDC, the total number of cells and average cell density were calculated based on occupied 100 × 100 µm spatial tiles, with tile area converted from µm² to mm² for reporting cell density. TDC were retained if they contained at least 100 cells and had a cell density of at least 500 cells/mm². A second level of filtering was then applied at the individual cell level, and cells with fewer than 10 Xenium UMI counts (nCount_Xenium > 10) were excluded.

Each sample (including all TDCs passing QC) was independently normalized using SCTransform, with all 196 genes in the targeted Xenium panel retained as variable features. Spatially informed clustering was performed using BANKSY with lambda = 0.2, M = 1, and k_geom = 15. PCA was performed on the BANKSY assay using 30 principal components. To account for sample- and section-specific technical variation, Harmony integration was performed using RunHarmony, grouped by spatial section and sample identifier. Harmony-corrected BANKSY embeddings were subsequently used for UMAP generation and nearest-neighbor graph construction. The first 20 Harmony dimensions were used for downstream analyses.

For cell-type annotation, the CosMx dataset was used as a reference for label transfer after restricting the reference to genes included in the Xenium 196-gene panel. Transfer anchors were identified using FindTransferAnchors, and predicted labels were assigned to Xenium cells using TransferData, thereby projecting CosMx-derived annotations onto the Xenium dataset.

### Spatial neighborhood and niche analysis

Spatial neighborhood analysis was performed using the CosMx label-transferred cell-type annotations. To prevent artificial neighborhoods spanning independent tissue regions, neighborhood composition was calculated independently within each tissue-density cluster. Within each tissue density cluster, the 20 nearest neighbors of every cell were identified from cell-centroid coordinates. A neighborhood-composition profile was generated for each cell by counting the number of neighboring cells belonging to each annotated cell type. Neighborhood-composition profiles from all tissue density cluster were subsequently combined into a single matrix. Cells with similar neighborhood-composition profiles were grouped using mini-batch K-means clustering. Multiple cluster numbers were evaluated, and five niches were retained based on the clarity and biological interpretability of cell-type-enriched neighborhood patterns. the cellular composition of each niche was summarized by averaging neighborhood-composition profiles across all cells assigned to the corresponding niche. The resulting niche-level composition matrix was standardized by Z-score transformation and visualized using a scaled heatmap.

## RESULTS

We examined a cohort (n = 24) of patients diagnosed with RA that either were naïve to therapy, or failed conventional Disease-Modifying Antirheumatic Drugs (DMARD) or biologic DMARD therapy and were candidates for ultrasound-guided synovial biopsy (**Table 1**). The average age was 52, the vast majority were female (>90%) and white (71%), 54% never smoked, and had an average disease duration of 90 months. Only 44% were RF+, while 39% were ACPA+. The average CDAI score was 23 and 46% of the patients were on DMARD therapy, while 42% were on biologic therapy. Approximately 67% of the patients had their wrist biopsied, while the other 33% underwent knee biopsy, all using an ultrasound-guided technique.

The biopsied synovial tissue was either immediately digested for single-cell analysis and/or placed in 4% neutral buffer formalin to be paraffin embedded for spatial transcriptomics. The single-cell suspension of the synovial tissue underwent fluorescent activated cell sorting (FACS) on the live CD45^+^ cells for processing on the 10X Chromium for Cellular Indexing of Transcriptomes and Epitopes by sequencing (CITE-seq). We clustered 37,459 CD45^+^ cells to yield 11 clusters and 5 major cell types (**Figure 1A**, **Supplemental Figure 1A-C**, and **Supplemental Table 4**). Myeloid cells were *CD68*+ and *LYZ*+, mast cells were *CPA3*^+^ and *KIT*^+^, T-cells were *IL7R*^+^ and *CD3*^+^, B-cells were *CD19*^+^ and *MS4A1*^+^, and plasma cells were *MZB1*^+^ (**Figure 1B**, **Supplemental Figure 1D**, **Supplemental Table 5**). Further, the gene expression pattern for the major cell types was confirmed by protein expression including CD3, CD8, CD4, CD19, CD20, FCERγ1α, CD11b and HLA-DR (**Figure 1C**). Across the subset of patients with both baseline and follow up (4-6 weeks later) biopsies, the majority of the CD45^+^ cells were T-cells, followed by myeloid cells; however, individual patients exhibited more variability (**Figure 1D-F**).

**Figure 1.**
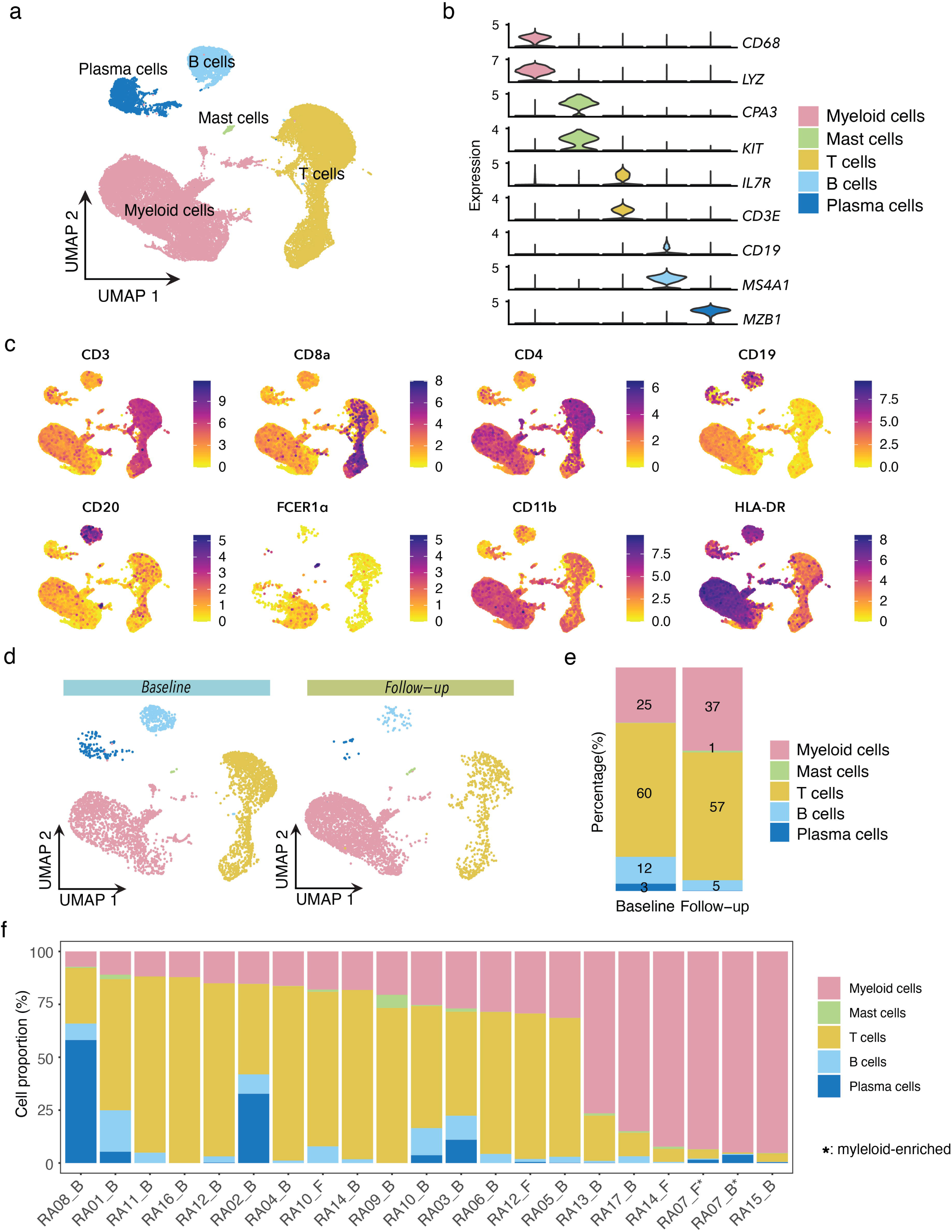
Single-cell landscape of rheumatoid arthritis synovial biopsies. (**a**) UMAP of n = 37,459 cells from 21 synovial biopsy samples, colored by major cell type. (**b**) Violin plots showing the expression of selected lineage markers across major cell populations. (**c**) ADT feature plots showing the expression of CD3, CD8a, CD4, CD19, CD20, FCER1α, CD11b, and HLA-DRA. (**d**) UMAPs showing cells from paired baseline and follow-up synovial biopsies. (**e**) Distribution of major cell populations in paired baseline and follow-up synovial biopsies. (**f**) Cell-type composition of individual synovial biopsy samples. Asterisks indicate myeloid-enriched biopsies.

We then extracted 16,761 myeloid cells across patients to identify macrophage subpopulations. Due to varying numbers of myeloid cells across samples, we first created a reference set using 5 patients (**Supplemental Figure 2A-B**). We then label-transferred the annotations and UMAP projection onto the remaining patient samples (**Figure 2A**) to yield 12 clusters; 8 macrophage subpopulations, conventional DCs (cDCs), plasmacytoid DCs (pDCs), osteoclast-like cells, and a proliferating population (**Figure 2A**-**B**, **Supplemental Table 6**). Macrophage subpopulations exhibited differential expression of *MERTK*, *TREM2*, *MRC1* and *CD163* as well as shared expression of *CD68*, *CSF1R*, *HLA-DRA*, and *CD14* (**Supplemental Figure 2C**). The individual subpopulations of macrophages were distinguished by expression of *IGF1* (Mac1), *CRIP2* (Mac2 and Mac3) and *TIMD4* (Mac2), *ISG15* (Mac4), *THBS1* (Mac5), *CCL4* (Mac6), *MT1 genes* (Mac7) and *S100A12* (Mac8) (**Figure 2B**, **Supplemental Table 6**). *CD1c*, *CLEC4C*, *MMP9* and *CDK1* defined the cDC, pDC, osteoclast-like and proliferating populations, respectively (**Figure 2B**). These subpopulations were further corroborated by surface expression of monocyte-derived (CD192, CD48) tissue-resident (CD206, Folrβ, CD1c) and common macrophage (MERTK, CD64, CD163, HLA-DR) markers (**Figure 2C** **and Supplemental Figure 2D**). Of note, we did not identify a specific *SPP1*^+^ population as Mac3, 4, 6 and 7 each expressed *SPP1*. We showed that the bulk of the macrophage populations were highly similar with the exception of Mac8 which exhibits a more monocyte-like profile (**Supplemental Figure 2E**). We then compared our 12 clusters to those proposed by Zhang et al [9] and Alivernini et al [12]. We identified similarities between CRIP2^+^TIMD4^+^ (Mac2) with M-2 *MERTK^+^S100A8^+^* and *TREM2^High^* and ISG15^+^ (Mac4) with M-6 *STAT1^+^CXCL10^+^* and *HLA^High^ISG15^+^* from Zhang et al. and Alivernini et al., respectively. We also found alignment for *S100A12*^+^ (Mac8) with M-7 *IL-1B^+^FCN1^+^* and *CD48*^high^*S100A12*^+^. Moreover, pDCs in our dataset resemble pDCs in Zhang et al. [9] and cDCs in our dataset resemble HLA^high^CLEC10A^high^ in Alivernini et al [12] (**Supplemental Figure 2F-G**). Although there is a variability between baseline and follow-up samples, we do not observe a consistent change in the four patients with paired samples (**Figure 2D-E**).

**Figure 2.**
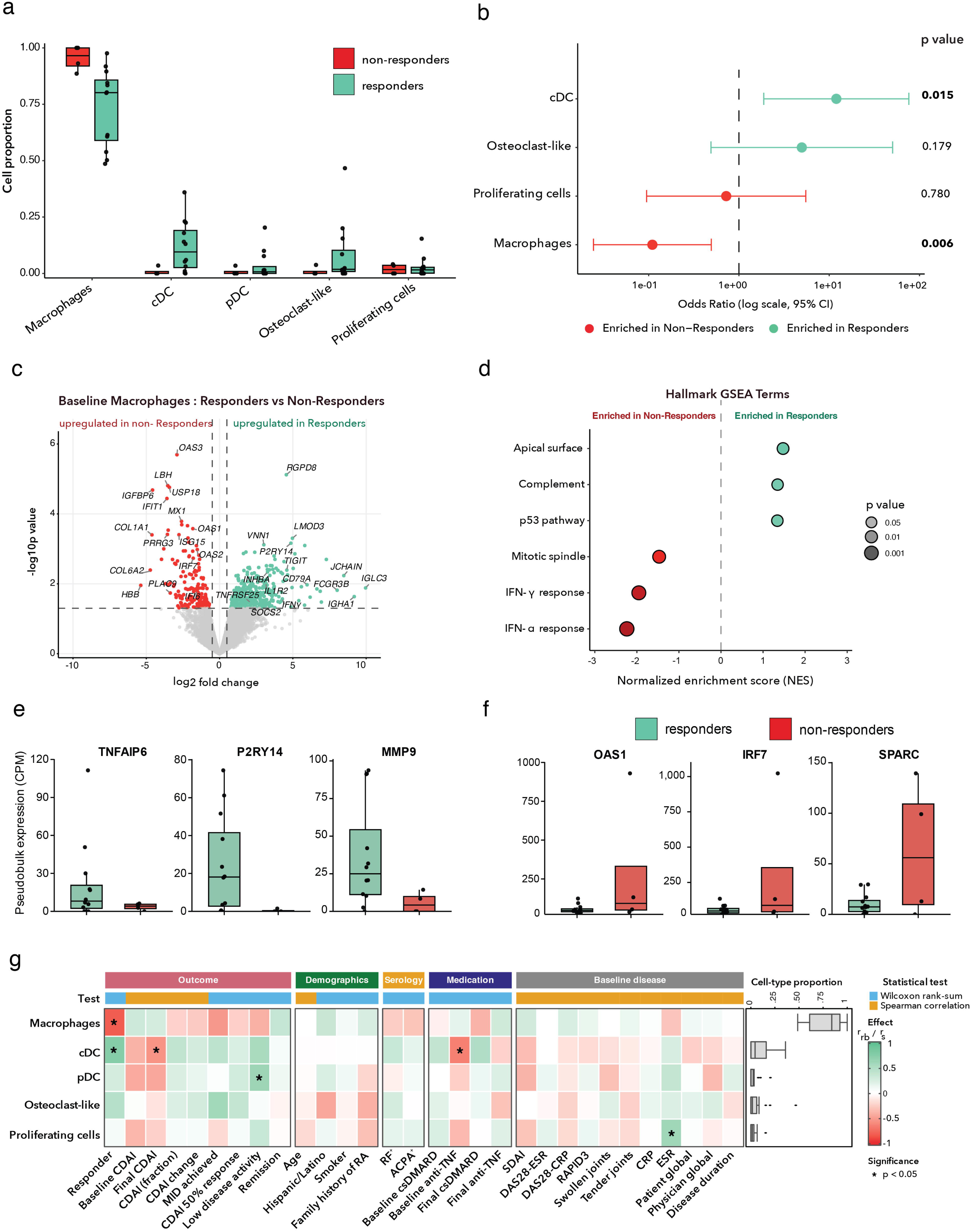
Myeloid cell heterogeneity in rheumatoid arthritis synovial biopsies. (**a**) UMAP of n = 16,761 myeloid cells showing unsupervised clustering into eight macrophage subsets, conventional dendritic cells (cDCs), plasmacytoid dendritic cells (pDCs), osteoclast-like cells and proliferating cells. (**b**) Violin plots showing the expression of selected marker genes defining individual myeloid cell populations. (**c**) ADT feature plots showing the expression of CD192, CD48, CD206 (MMR), Folate Receptor β, and CD1c. (**d**) UMAPs showing the distribution of myeloid cell populations in paired baseline and follow-up synovial biopsies. (**e**) Proportions of myeloid cell populations in paired baseline and follow-up synovial biopsies. Colored points represent matched patient samples

Patients who achieved 20% improvement in CDAI score (CDAI-20) were considered responders and those that failed to achieve CDAI-20 were non-responders [42, 43]. As expected, there were significant differences in final CDAI score, CDAI change, fractional CDAI improvement, and MID achieved between these groups (**Supplemental Table 7**). We compared the percent of total macrophages, cDCs, osteoclast-like cells, pDCs and proliferating cells at baseline from responders vs. non-responders. We observed a significant odds ratio indicating a decrease (OR = 0.16, p = 0.006) in the proportion of macrophages and an increase (OR = 12.0, p = 0.015) in the proportion of cDCs in patients who responded compared to those who failed to respond (**Figure 3A-B**, **Supplemental Table 8**). We then examined the differential gene expression between responders and non-responders in macrophages. We detected increased *TNFAIP6*, *PLAUR, MMP9, CXCL1, BMP2,* and *P2RY14* but reduced *OAS1, IFIT1, MX1, ISG15, SPARC* and *IRF7* expression in responders (**Figure 3C-F**, **Supplemental Table 9**). These genes corresponded to an upregulation of complement and p53 pathways in responders and enrichment of IFN pathway in non-responders (**Figure 3D**, **Supplemental Table 10**). The cDC population expressed increased *VEGFβ, HK2, TLR3/7, MIF, TGFα, VCAN, IFIT1* and *OAS2* in non-responders (**Supplemental Figure 3A**, **Supplemental Table 9**), which were associated with an enrichment of IFN response, hypoxia, angiogenesis and glycolysis pathways (**Supplemental Figure 3B**, **Supplemental Table 9-10**). The macrophage and DC populations were associated with response status, while DC proportions were also negatively correlated with final CDAI score and decreased in patients undergoing baseline anti-TNF therapy. pDCs were associated with low disease activity (**Figure 3G**).

**Figure 3.**
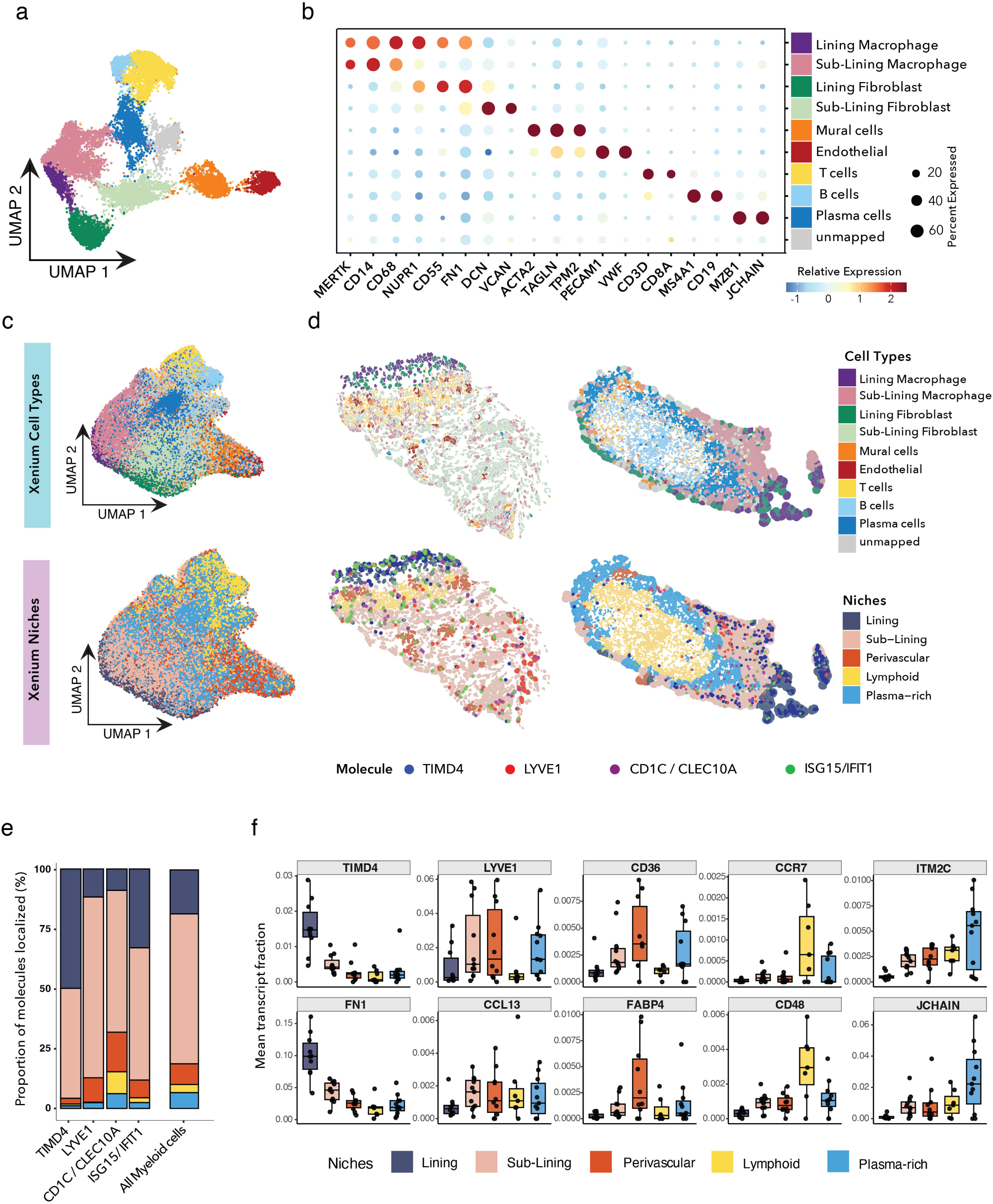
Baseline myeloid cell populations associated with clinical response. (**a**) Sample-level proportions of major myeloid cell populations in baseline synovial biopsies from responders (n = 12) and non-responders (n = 4). **(b)** Association of baseline major myeloid cell populations with clinical response estimated using mixed-effects association testing (MASC). Points indicate odds ratios, error bars represent 95% confidence intervals, and nominal p-values are shown. (**c**) Volcano plot showing differentially expressed genes from pseudobulk differential expression analysis of baseline macrophages comparing responders and non-responders. (**d**) Hallmark gene set enrichment analysis of pseudobulk differentially expressed genes from baseline macrophages. Dot color and size indicates nominal p-value and the x axis shows the normalized enrichment score (NES). (**e,f**) Pseudobulk expression (CPM) of selected differentially expressed genes in baseline macrophages from responders (**e**) and non-responders (**f**). **(g)** Associations proportions of cell populations with clinical variables using the Wilcoxon rank-sum test for binary variables and Spearman’s rank correlation for continuous variables. Associations were visualized using heatmaps displaying the corresponding effect sizes (rank biserial correlation r_rb_ or Spearman’s coefficient r_s_), with nominal significance indicated at p < 0.05.

To further investigate patterns of response-based gene expression across macrophages, we independently clustered responder-specific (K = 4) and non-responder-specific (K = 3) genes using K-means clustering of the differentially expressed genes from pseudobulk macrophages (**Supplemental Figure 4**). We found that Mac1, Mac2, Mac3 and Mac5 were mainly responsible for the increase in genes in Cluster 1 associated with *retinoid metabolic process* and including *ALDH1A1*, *GDF15, and PTGDS*. Genes that represent *inflammatory response*, such as *IL1R2* and *MMP9,* were higher in Mac5, Mac6 and Mac8 in responders. *Chemotaxis* pathway genes including *TREM1, FCGR3B* and *TNFAIP6* was elevated in Mac4, Mac5 and Mac 8 subpopulations in responders. *IL-1B production* was driven by Mac5, Mac7 and Mac8 in responder patients (**Supplemental Figure 4A-C**). We observed that Mac1, Mac2, Mac5 and Mac8 were responsible for *post-transcriptional regulation of gene expression* in non-responders. Mac3 and Mac4 accounted for the upregulation of *skeletal system development* and *response to virus* pathways, respectively, in non-responders (**Supplemental Figure 4B-D, Supplementary Table 11-12**).

We then examined whether the proportion of individual macrophage subpopulations correlated with disease outcomes. We found that the proportions of the *ISG15^+^* (p = 0.005) and *THBS1^+^* (p = 0.047) macrophages were enriched in non-responders (**Figure 4A-B, Supplemental Table 8**). *CDKN1A, CXCL10, IL-1β, IRF7, CD36* and *SMAD4/5* expression were increased in *ISG15+* non-responder macrophage population, which coincided with enrichment of p53, IFN, hypoxia and mTOR signaling pathways (**Figure 4C-D, Supplemental Tables 9-10**). Upregulation of NDUFA6, COX17, CCT3, and RPS2 led to the enrichment of the oxidative phosphorylation and myc-target pathways in responders in *THBS1^+^* macrophages. The *THBS1^+^* macrophage subpopulation also exhibited increased expression of *IFI44*, *OAS3*, and *IFI27* which are associated with the interferon pathways in non-responders (**Figure 4E-F, Supplemental Tables 9-10**). Proportions of *CRIP2^+^TIMD4^-^*(Mac3) macrophages were decreased in the Latino/Hispanic population. The *ISG15^+^* macrophages were negatively associated with response to therapy and the minimally important difference (MID) in CDAI [44]. The *THBS1^+^* macrophages were negatively correlated with change in CDAI, reduced in RF^+^ patients, and positively correlated with age.

**Figure 4.**
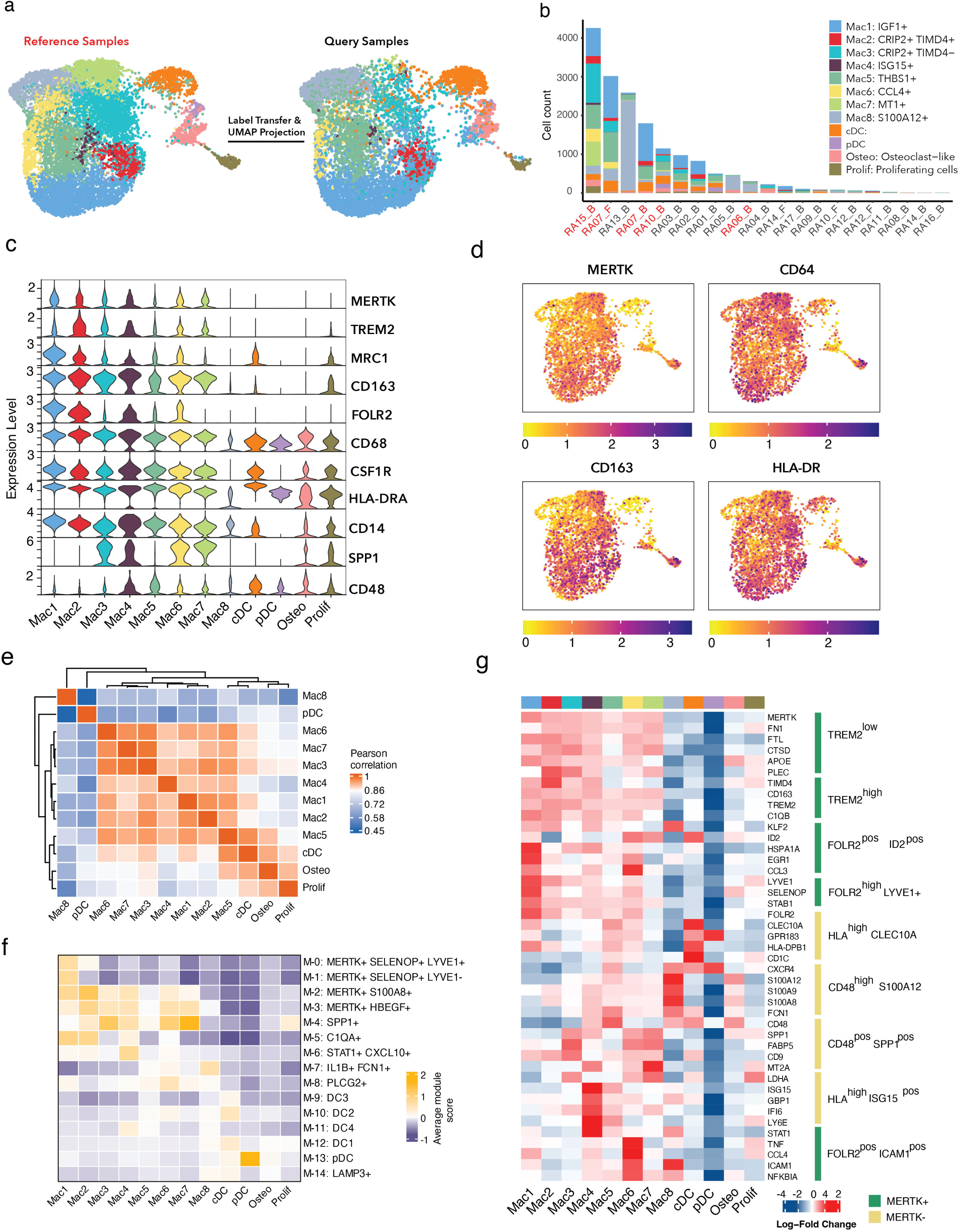
Baseline macrophage subsets associated with clinical response. (**a**) Sample-level proportions of macrophage subpopulations in baseline synovial biopsies from responders (n = 12) and non-responders (n = 4). (**b**) Association of baseline myeloid cell populations with clinical response estimated using mixed-effects association testing (MASC). Points indicate odds ratios, error bars represent 95% confidence intervals, and p-values are shown. (**c**) Volcano plot showing pseudobulk differential gene expression between Mac4 (*ISG15⁺*) macrophages from responders and non-responders. (**d**) Hallmark gene set enrichment analysis of the Mac4 differential expression results. Dot color and size indicates nominal p-value and the x axis shows the normalized enrichment score (NES). (**e**) Volcano plot showing pseudobulk differential gene expression between Mac5 (*THBS1⁺*) macrophages from responders and non-responders. (**f**) Hallmark gene set enrichment analysis of the Mac5 differential expression results. Dot color and size indicates nominal p-value and the x axis shows the normalized enrichment score (NES). **(g)** Associations proportions of macrophage subpopulations with clinical variables using the Wilcoxon rank-sum test for binary variables and Spearman’s rank correlation for continuous variables. Associations were visualized using heatmaps displaying the corresponding effect sizes (rank biserial correlation r_rb_ or Spearman’s coefficient r_s_), with nominal significance indicated at p < 0.05.

We performed spatial transcriptomics on synovial biopsy tissue using the 1000 gene discovery panel on the CosMx Spatial Molecular Imager. We identified 15,874 cells that denoted macrophages (lining and sublining), fibroblasts (lining and sublining), mural cells, endothelial cells, and lymphocyte based on previous reports [9, 12] (**Figure 5A-B, Supplemental Figure 5A-B**). We then used the CosMx dataset as the reference to annotate synovial biopsy tissue profiled by Xenium (n = 10) using a custom 196-gene panel designed to distinguish macrophage subpopulations (**Supplemental Figure 5C**). Although enriched for macrophage-associated genes, the panel also includes markers of lymphocytes, fibroblasts, and endothelial cells. We examined 93,028 cells and were able to define similar cell types (**Figure 5C**) compared to CosMx (**Supplemental Figure 5D**), as well as niches that define lining, sublining, perivascular, lymphoid and plasma cell-rich regions (**Figure 5D, Supplemental Figure 5E**). The 196 custom panel contained *ISG15* but did not have *THBS1*. *TIMD4* (highest expression in Mac2) and *ISG15/IFIT1* molecules were enriched in macrophages from the synovial lining suggesting that these populations are localized there. In contrast, *LYVE1* (highest expression in Mac1) and *CD1c*/*CLEC10A* molecules were mainly localized to macrophages in the sublining niches (**Figure 5E-F**). Since *THBS1^+^* macrophages highly express CD48 (**Supplemental Figure 2G**), which is found in macrophages in the lymphoid niche of the sublining (**Figure 3**), we can infer that the *THBS1^+^* macrophages are localized to the sublining. Collectively, these data suggest that proportions of macrophage subpopulations are associated with response to therapy in RA patients and are localized to both the lining and sublining areas.

**Figure 5.**
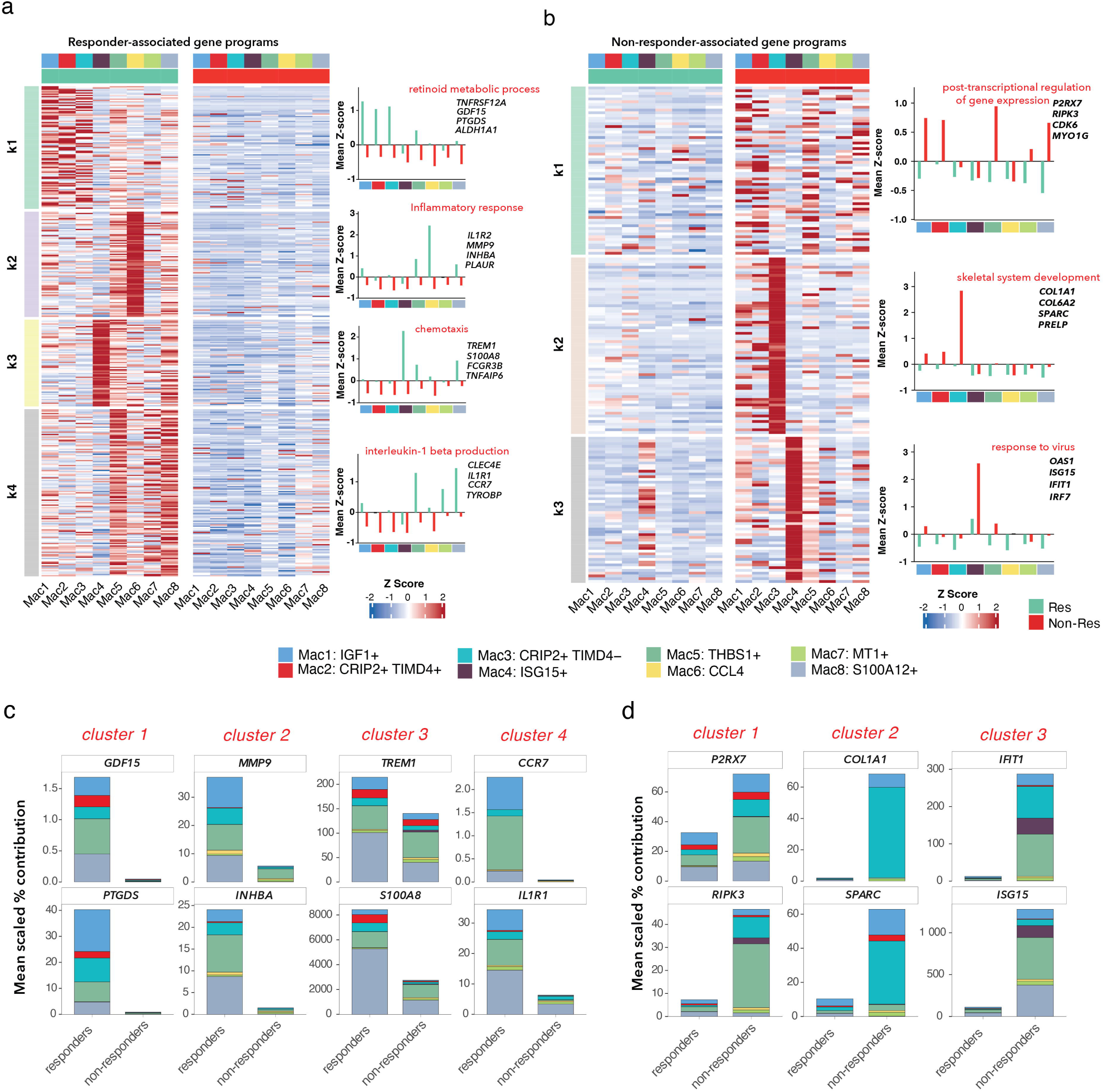
Spatial mapping of synovial cell states and myeloid niches. (**a**) UMAP of 15874 cells from one CosMx synovial biopsy, colored by annotated cell type. (**b**) Dot plot showing the expression of selected marker genes across annotated CosMx cell types. Dot size indicates the percentage of expressing cells and color indicates relative expression. (**c**) UMAP of 93028 cells from 11 synovial biopsy samples, colored by transferred cell-type labels (top) and spatial niche annotations (bottom). (**d**) Spatial distribution of annotated Xenium cell types (top) and niches overlaid with molecules (*TIMD4*, *LYVE1*, *CD1C/CLEC10A*, and *ISG15/IFIT1*) in macrophages (bottom). (**e**) Proportion of molecules localized to in macrophages across Xenium spatial niches. (**f**) Fraction of transcripts for selected genes out of total transcripts detected in the cell averaged across all cells in the given spatial niches for each Xenium sample.

## DISCUSSION

Changes in macrophage numbers have been linked to response to therapy in patients with RA [5, 6]. We were one of the first to show that a subpopulation of synovial macrophages (CD206^+^) in RA express six different transcriptional modules, which are associated with a therapy and disease severity [7]. Here, we present a prospective, longitudinal study of RA patients who had ultrasound-guided synovial biopsies for single-cell RNA seq and for spatial transcriptomics, prior to change in therapy and monitored over time. We observed that the proportion of total macrophages is reduced in RA patients who respond to therapy. Further, we identified that *ISG15^+^* and *THBS1^+^* macrophage subpopulations correlate with response to therapy as indicated CDAI-20. Moreover, the *ISG15^+^* and *THBS1^+^* populations appear to localize preferentially to the lining and sublining tissue, respectively. This study is the first to link transcriptionally defined synovial macrophage subpopulations, profiled prospectively before a change in therapy, to subsequent therapeutic response.

Few studies have utilized multi-omic analyses of synovial tissue in RA patients pre- and post-treatment. The Pathobiology of Early Arthritis Cohort (PEAC) performed ultrasound guided synovial biopsies in early RA diagnosed patients prior to any therapy including steroids. Synovial tissue was analyzed by histology and bulk RNA seq as well as peripheral blood mononuclear (PBMCs) by bulk RNA seq followed by deconvolution analysis [15]. The authors identified 3 histologic patterns in synovial tissue; lympho-myeloid, diffuse myeloid, and pauci-immune fibroid, although this classification was insufficient to predict response to therapy. They further showed that the transcriptional signatures of whole peripheral blood were not associated with clinical response. However, they identified transcriptional modules from whole synovium that correlates with response to DMARD therapy and radiographic progression [15, 16]. In the R4RA trial, the goal was to compare tocilizumab vs rituximab in anti-TNF non-responder RA patients [17]. The authors showed that histologic analysis of the synovial biopsy prior to change in therapy was inadequate to predict preferential response to a particular therapy similar to the PEAC study. However, when tissue samples were classified as B-cell poor or B-cell rich by transcriptomic analysis, tocilizumab was superior to rituximab in the B-cell poor group based on achieving a 50% change in CDAI or a change in CDAI major treatment response [17]. No significant differences in response rates were found in the B-cell rich population between tocilizumab- and rituximab-treated patients [17]. Additional analyses identified humoral immune signatures associated with treatment response, whereas a stromal/fibroblast transcriptional signature was associated with failure to respond to both tocilizumab and rituximab [18]. The stratification of biological therapies by pathobiology in biologic-naïve patients with rheumatoid arthritis (STRAP and STRAP-EU) was constructed to evaluate B-cell rich and B-cell poor, DMARD non-responders who were switching to rituximab, tocilizumab or etanercept. The classification based on B-cell composition was not sufficient to predict response to any therapy, especially anti-B-cell therapy such as rituximab [19]. However, a 524 gene panel may predict response to rituximab, tocilizumab or etanercept using bulk transcriptomic analyses and machine learning models [20], although this has not been verified by others. One of the central weaknesses of PEAC, R4RA and STRAP was the utilization of bulk RNA seq analysis and deconvolution techniques, which are not optimal for analyzing the cellular proportions and transcriptional signatures at cell-type or cell-state level [47, 48]. Nonetheless, these data support that notion that transcriptomic analysis has the potential to unlock predictive models for therapeutic response in RA patients. In our prospective study, we identified the proportion of macrophages as a whole population correlates with response to therapy. Further, we identified two subpopulations of macrophages (*ISG15^+^* and *THBS1^+^*) whose proportions correlate with response to therapy. Future studies will be required to validate the two subpopulations of macrophages as well as determine whether they are associated with individual therapies.

Over the past several years, single cell transcriptomics have been applied to deciphering the heterogeneity of the cellular populations within the synovial tissue. Our data showed a lower total proportion of macrophages in the responder population, although we did not determine whether the reduction was in the synovial lining or sublining macrophages. The Accelerating Medicines Partnership initially identified 4 synovial macrophage populations [8], which were further resolved into 9 populations in a subsequent study [9]. The latter study also defined cell type abundance phenotypes (CTAPs) based on the relative composition of multiple cell types within synovial tissue cross-sectional study. None of the CTAP were associated with clinical metrics based on their cohort [9]. However, the authors also applied CTAPs derived from the single cell studies to a bulk RNA seq dataset from the R4RA trial [9]. They demonstrated that the patients with the fibroblast CTAP displayed the poorest response to either treatment [9]. Alivernini et al also performed a cross-sectional study on RA patients undergoing synovial biopsy followed by scRNA seq but was able to clinically stratify patients based on their stage of disease or response to therapy [12]. While Alivernini et al. did not examine DCs in their dataset [12], MacDonald et al. identified subpopulations of DCs and their localization and interactions with macrophages and lymphocytes [45]. They observed that the proportion of total DCs (except pDCs) is markedly increased in the responders compared to non-responders. Future studies will examine the subpopulations of DCs to determine which ones contribute to the response or failure to respond to therapy.

Our patients’ therapy ranged from methotrexate, anti-TNF, anti-IL-6 to JAK inhibitors following biopsy. There is no specific therapy linked to the non-responder patients as half were treated with an anti-TNF, while the others were treated with a JAK inhibitor or anti-IL6 inhibitor. Interestingly, pathways associated with type 1 and type 2 interferon were enriched in non-responders across all macrophages as well as in subpopulations of macrophages. While anifrolumab, an inhibitor of the type I interferon pathway is approved for patients with systemic lupus erythematosus (SLE) [46] and has been successful at treating rash and inflammatory arthritis [47], there are only few studies that examined anifrolumab for RA. In small pilot study, clinical efficacy was observed in patients treated with anifrolumab [48], however, there are no planned or published studies for an extended trial in RA using anifrolumab. Nonetheless, our data suggest that for non-responder patients, anifrolumab may be a viable candidate if the patients display a type I interferon response. Hypoxia and angiogenesis were also enriched in non-responder macrophage subpopulations. In the synovium, the efficiency of oxygen supply is low, which leads to a hypoxic gradient and greatly affects the behavior as well as function of the synoviocytes [49]. Thus, new therapeutics that augment angiogenesis and reduce hypoxia could potentially also be advantageous for the non-responder populations.

There are limitations to the study including the number of patients, mixed therapies, the few numbers of patients undergoing synovial biopsy at 4-6 weeks post-treatment and the use of a macrophage specific gene array for the Xenium spatial imaging, which precluded the corresponding analysis of all cell populations in the synovium. Future studies will be required to examine patients prior to a specific therapy similar to the STRAP and R4RA trial but utilize single cell analysis or full transcriptome wide spatial sequencing. Furthermore, an increase in the number of patients undergoing a second biopsy would be ideal but was not a possibility for this trial as COVID limited the return of patients for a second biopsy. Nonetheless, the findings from this novel, prospective study in RA establish a role for synovial macrophage subpopulations as indicators of therapeutic response.

## Data Availability

All data produced in the present study will be made available upon study publication in a peer-reviewed journal.

## ACKNOWLEDGMENTS

We would like to thank the Northwestern University Lurie Cancer Center Flow Cytometry Core Facility, which is supported by NCI Cancer Center Support Grant P30 CA060553 awarded to the Robert H. Lurie Comprehensive Cancer Center. This research was supported in part through the computational resources and staff contributions provided by the Genomics Compute Cluster, which is jointly supported by the Feinberg School of Medicine, the Office of the Provost, the Office for Research and Northwestern Information Technology. The Genomics Compute Cluster is part of Quest, Northwestern University’s high-performance computing facility, with the purpose to advance research in genomics. MD was supported by the National Scleroderma Foundation (New Investigator Award) and the Rheumatology Research Foundation (Investigator Bridge Award). SDD was supported by the Arthritis National Research Foundation (Postdoctoral Research Fellowship), the Brinson Foundation, and AR085530. CMC was supported by the Lupus Research Alliance (Novel Research Grant), the Rheumatology Research Foundation (Innovative Research Grant) and AI170938. HMM was supported by KL2 TR001424, HL134375S1, and AR007611. DRW was supported funding from Arthritis National Research Foundation, American Federation for Aging Research/Hevolution, American Heart Association (18CDA34110224) and NIH grant AI163742 and AR080513. HP was supported by AR080513, AR075423, HL169188, CA060553, AR085530, the Precision Medicine Fund, and the Mabel Green Myers Professorship. The authors declare no competing financial interests.

## FIGURE LEGENDS

**Supplemental Figure 1.**
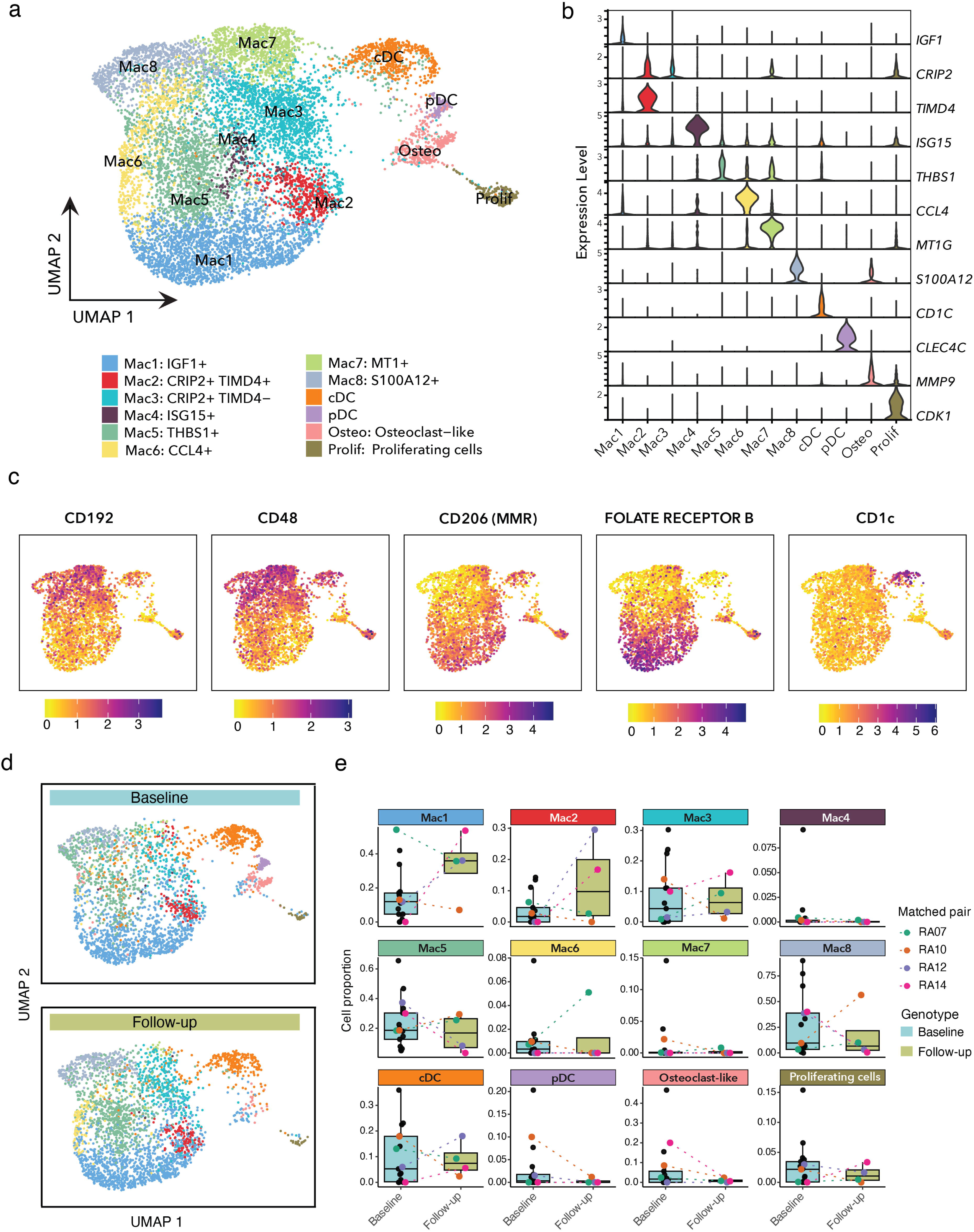
Quality control and annotation of synovial biopsy single-cell transcriptomes. (**a**) Violin plots showing detected genes per cell, UMI counts per cell, mitochondrial transcript percentage and ribosomal transcript percentage across synovial biopsy samples. (**b**) UMAP of the integrated dataset showing unsupervised cell clusters. (**c**) Heatmap showing the top de novo marker genes for each unsupervised cluster. (**d**) Dot plot showing canonical lineage-marker expression across unsupervised clusters. Dot size represents the percentage of cells expressing each gene, and color indicates relative expression.

**Supplemental Figure 2.**
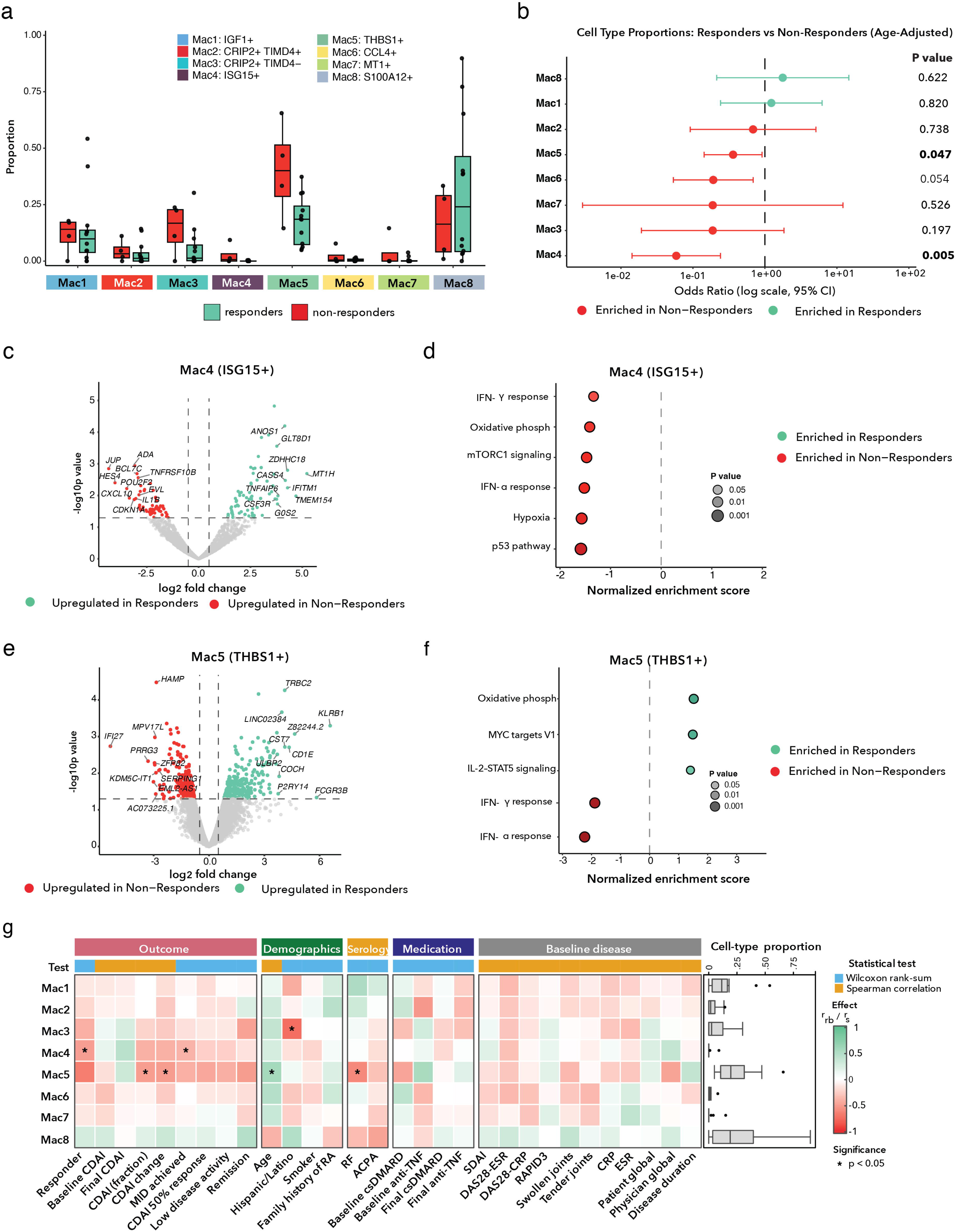
Annotation and validation of synovial myeloid cell populations. (**a**) Label transfer and UMAP projection of query samples using five reference synovial biopsy samples with manually annotated myeloid cell populations. (**b**) Number of cells annotated by myeloid cell population across individual synovial bioly samples following label transfer. Red sample IDs indicate reference samples. (**c**) Violin plots showing the expression of selected myeloid marker genes across annotated myeloid cell populations. (**d**) Surface protein (ADT) feature plots showing the expression of MERTK, CD64, CD163 and HLA-DR across annotated myeloid cell populations. **(e)** Pearson correlation of the 12 myeloid populations. (**f**) Heatmap showing module scores of synovial myeloid subset signatures from Zhang et al. [9] across annotated myeloid cell populations. (**g)** Heatmap showing the log-fold change of Alivernini et al. [12] synovial myeloid subset signature genes across annotated myeloid cell populations.

**Supplemental Figure 3.**
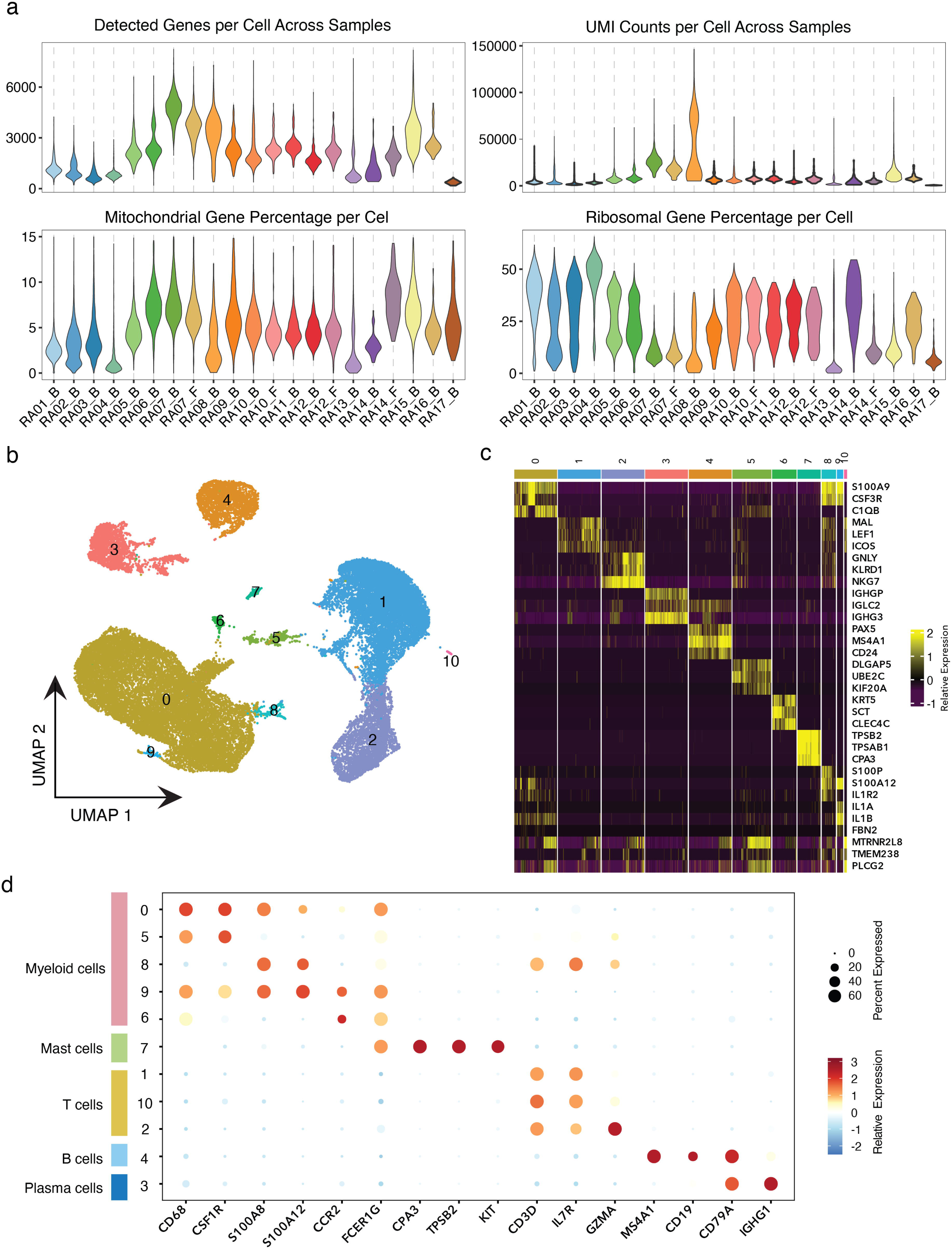
Baseline cDCs associated with clinical response. (**a**) Volcano plot showing pseudobulk differential gene expression in baseline conventional dendritic cells from responders and non-responders. (**b**) Hallmark gene set enrichment analysis of the conventional dendritic cell differential expression results. Dot color and size indicates the nominal p-value and the x axis shows the normalized enrichment score (NES).

**Supplemental Figure 4.**
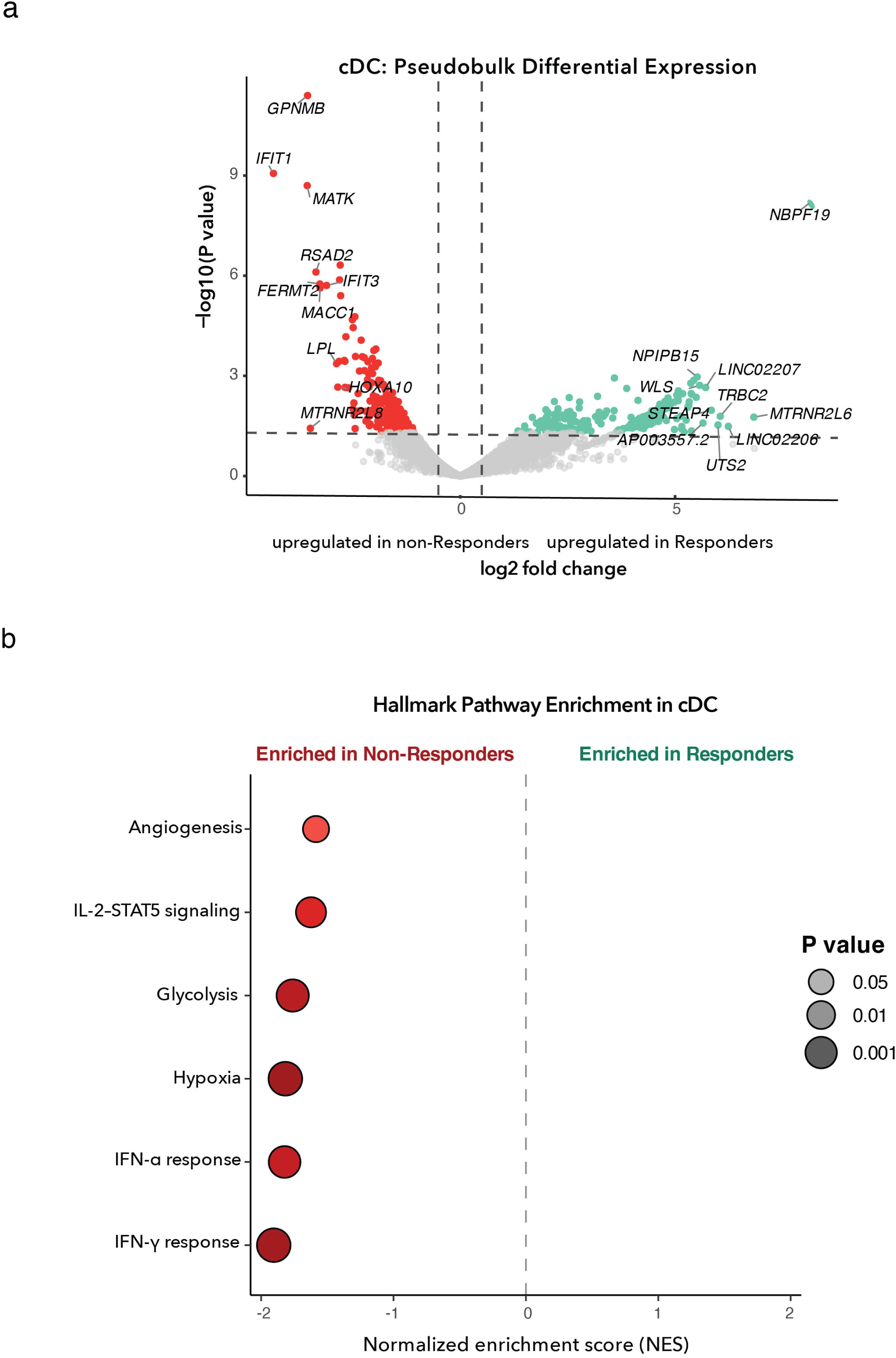
Baseline transcriptional programs associated with clinical response in macrophages. (**a**) K-means clustering of genes upregulated in responder macrophages. Heatmaps show row-scaled sample-level mean normalized expression across Mac1–Mac8 populations in responders and non-responders, with genes partitioned into four clusters. Adjacent plots show the mean Z-score profile for each cluster. (**b**) K-means clustering of genes upregulated in non-responder macrophages. Heatmaps show row-scaled sample-level mean normalized expression across Mac1–Mac8 populations in responders and non-responders, with genes partitioned into three clusters. Adjacent plots show the mean Z-score profile for each cluster. (**c**) Relative contribution of Mac1–Mac8 populations to the expression of selected responder-associated genes. Stacked bars show the mean per-sample percentage contribution of each macrophage population scaled by the mean pseudobulk expression within each response group. (**d**) Relative contribution of Mac1–Mac8 populations to the expression of selected non-responder-associated genes. Stacked bars show the mean per-sample percentage contribution of each macrophage population scaled by the mean pseudobulk expression within each response group.

**Supplemental Figure 5.**
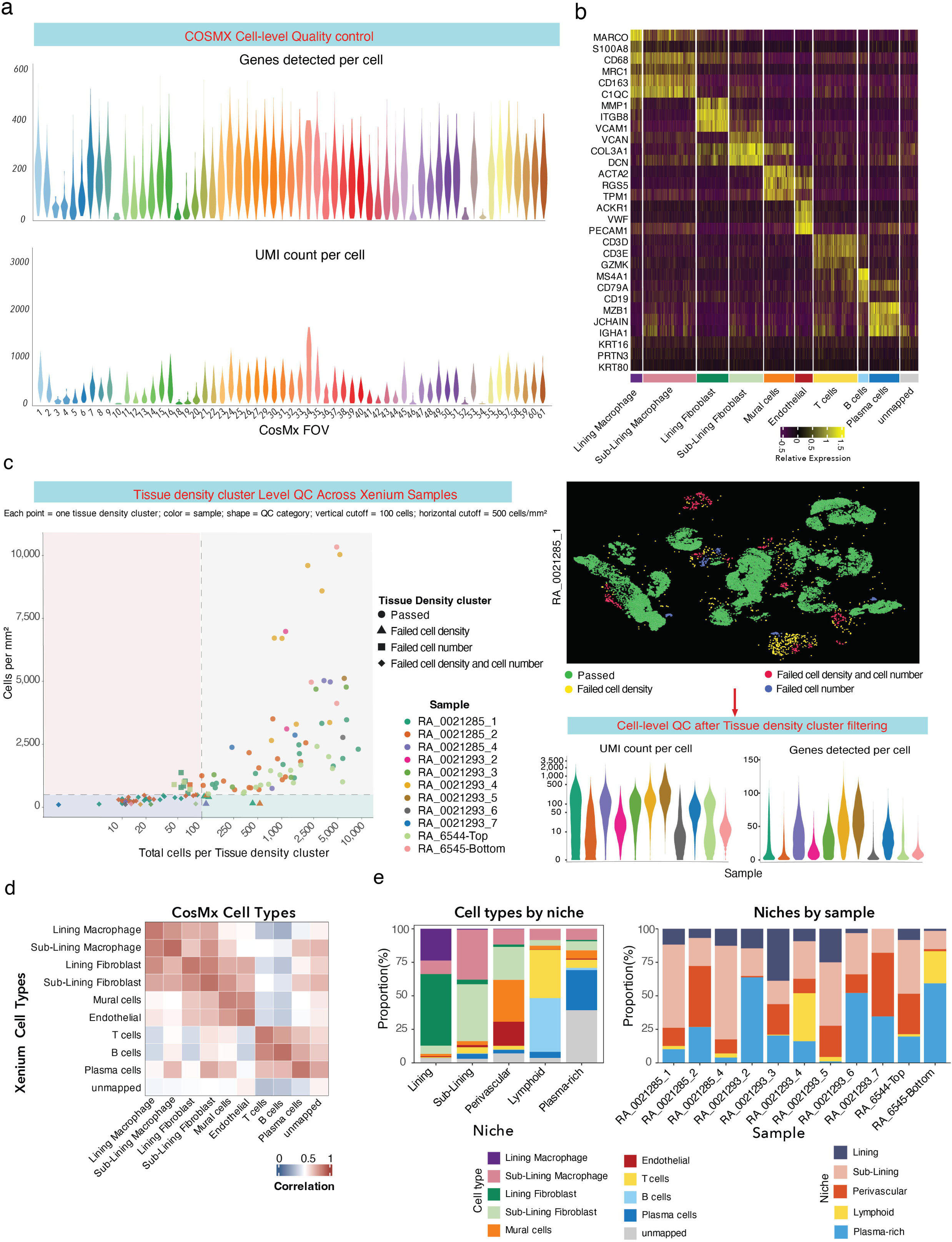
Quality control and cross-platform validation of CosMx and Xenium spatial transcriptomic datasets. (**a**) Cell-level quality control metrics across all CosMx fields of view showing the number of genes detected and UMI counts per cell. (**b**) Heatmap showing the expression of de novo marker genes across annotated CosMx cell types. (**c**) Xenium quality-control workflow. Left, quality assessment of DBSCAN-defined tissue chunks across all Xenium samples. Top right, representative tissue chunk classification in a representative Xenium sample. Bottom right, cell-level quality-control metrics following tissue density filtering. (**d**) Correlation of expression for shared genes between the CosMx reference and Xenium dataset following label transfer. (**e**) Composition of annotated cell types across Xenium spatial niches (left) and of niches across Xenium samples (right).

